# Effects of Pod-Based Electronic Cigarette Use on Vascular Health and Relation to Volatile Organic Compound Exposure in Young Adults

**DOI:** 10.1101/2022.12.16.22283590

**Authors:** Sana Majid, Jessica L. Fetterman, Robert M. Weisbrod, Leili Behrooz, Andrew Stokes, Michael J. Blaha, Daniel Conklin, Sanjay Srivastava, Rose Marie Robertson, Aruni Bhatnagar, Rachel J. Keith, Naomi M. Hamburg

**Author notes:** These authors contributed equally (Rachel J. Keith, Naomi M. Hamburg). Corresponding Author: Naomi M. Hamburg, MD, Whitaker Cardiovascular Institute, Boston University School of Medicine, 72 East Concord St, Boston, MA 02118.

## Abstract

**Background:** Pod-based electronic cigarettes (e-cigarettes) use that contain nicotine salts is frequent among youth and young adults; thus, we compared the vascular health effects of pod-based e-cigarette use to combustible cigarette use.

**Methods and Results:** We performed a two center observational, cross-sectional study of healthy adults recruited from the community (aged 18-45, N=106) in 3 groups: pod-based e-cigarette users (N=48); combustible cigarette users (N=21); and tobacco nonusers (N=37) and assessed the acute (following structured use) and chronic (resting state after 6 hour tobacco abstinence) effects of pod-based e-cigarette use on endothelial function (brachial artery flow-mediated dilation), blood pressure, and heart rate. Among the pod-based e-cigarette users, 64% were exclusive users including 37% who had never used combustible cigarettes. Pod-based e-cigarette users and combustible cigarette users had higher systolic blood pressure compared to non-users (121±11mmHg, 121±13mmHg, 112±10 mmHg, P=0.0004). Structured pod-based e-cigarette use acutely decreased flow-mediated dilation (−3.2±2.7%), raised systolic and diastolic blood pressure (6±8mmHg, 4±5mmHg) and heart rate (5±7bpm), similar to combustible cigarette use (−2.6±2.6%, 9±8mmHg, 6±5mmHg, 6±6bpm P=0.83, 0.3, 0.4, 0.56 vs pod-based), and to a greater extent than nonuse (0.3±4.1%, 0.7±5mmHg, 0.3±3mmHg, -3±4bpm, P=1.0x10^−7^, 0.002, 0.003, 2.6x10^−7^). Differences remained robust in models adjusted for age, sex, and race. The effect of pod-based cigarette use was similar in adults who had never used combustible cigarettes. Levels of acrolein, acrylamide, acrylonitrile, and crotonaldehyde were associated with the changes in vascular health measures.

**Conclusions:** Overall, our findings suggest that pod-based e-cigarette use has acute and chronic vascular effects in healthy young adults including those who never used combustible cigarettes. Select metabolites derived from volatile organic compounds were associated with the vascular changes suggesting relevance to vascular health.

## INTRODUCTION

Electronic cigarettes (e-cigarettes) are popular tobacco products, particularly among youth and young adults.^1^ E-cigarette products are marketed as a safer alternative to combustible cigarettes, contributing to youth use.^2^. Many young e-cigarette users have never smoked combustible cigarettes, raising concerns of nicotine addiction exclusively through the use of e-cigarettes.^3, 4^ The rise in e-cigarette use among youth corresponded with the popularity of a new type of e-cigarette - pod-based devices, including JUUL.^5^

Since their introduction in 2015, pod-based e-cigarettes have become the dominant e-cigarette product sold in the United States, with JUUL accounting for the majority of the current market share.^5^ In contrast to older e-cigarette models, pod-based e-cigarettes utilize nicotine salts, which are more soluble in the propylene glycol and vegetable glycerin solvents than free base nicotine, and thereby enable greater nicotine absorption.^6^ The unique constitutents and mode of nicotine delivery raise the possibility that the health effects of pod-based e-cigarettes may be different from those associated with the use of earlier generations of e-cigarettes. This seems particularly plausible given that previous research with earlier generation devices showed that the acute vascular changes elicited by e-cigarette use were, in part, attributable to nicotine.^7^ Moreover, pod-based devices may generate different levels of reactive volatile organic compounds (VOCs), which could contribute to vascular dysfunction and injury.

The potential toxicity of pod-based devices is highlighted in a recent study using a rat model, showing that JUUL exposure leads to acute deterioration of endothelium-dependent vasodilation to a similar extent as was observed with combustible cigarette exposure.^8^ However, little is known in regard to the health effects of pod use, particularly in young adults, who are the most frequent users of these devices. The current study was therefore designed to examine the chronic and acute vascular effects of pod-based e-cigarette use in young, healthy adults compared to nonuse and combustible cigarette use and to evaluate the association between exposure to VOCs exposure and vascular health.

## METHODS

Primary data supporting these finding are available from the American Heart Association upon request.

### Study Participants

To examine acute vascular effects of e-cigarettes, we recruited 18-45 year old men and women from the community, who were without established cardiovascular disease or risk factors (dyslipidemia, hypertension, diabetes) at Boston University Medical School and University of Louisville, as previously described.^9^ We assessed self-reported tobacco use with a modified version of the National Health Interview Survey on Tobacco Use. Additional questions were added for e-cigarettes and vape devices and were harmonized, when possible, with the PhenX toolkit.^10^ Self-reported tobacco product use was used to assign participants into three categories: 1. Pod-based e-cigarette use by current use for the past 30 days, with use on at least five days per week; 2. Combustible cigarette users by the use of cigarettes for the past 30 days, with at least five days of use per week, a minimum of five cigarettes per day, without the use of e-cigarettes; and 3. Non-users of tobacco were defined as having less than 100 lifetime uses of a tobacco product and no use in the past 30 days. Pod-based e-cigarette users were comprised of dual users (ongoing combustible cigarette use), exclusive users who were former combustible cigarette users (at least 3 months of abstinence from combustible cigarette) and exclusive users who were never combustible cigarette users. All participants were instructed to refrain from tobacco product use at least 6 hours prior to the study visit and fast for 8 hours. All participants gave written informed consent and all study protocols were approved by the University of Louisville and the Boston University Institutional Review Boards.

### Acute Use Protocol

We observed participants before and after a structured use of their usual tobacco product. Tobacco product users were instructed to bring their usual tobacco product to the study visit. Pod-based e-cigarette users (both dual and exclusive users) were instructed to bring the type of pod they used most frequently. Combustible cigarette users smoked their usual cigarette brand. Pod-based e-cigarette users were asked to take a minimum of one puff per min and no more than two puffs per min for a total of 10 min. Combustible cigarette users smoked one cigarette in 10 min. Non-users were instructed to breath through a straw for 10 minutes. Given the lack of available studies of pod-based e-cigarette use topography,^11^ the study protocol was based on prior studies of e-cigarette topography. Controls followed a similar timing schedule of testing, but were not exposed to any tobacco product. The study did not constitute a clinical trial as individuals used their own products.

### Vascular Function Testing

Resting blood pressure and heart rate were recorded in triplicate after 10 min in recumbent position, using an automated blood pressure cuff (Omron, Kyoto, Japan). Non-invasive vascular function was measured as previously described to assess baseline brachial artery diameter and flow-mediated dilation.^9, 12^ Brachial artery diameter was measured at baseline and after a 5 min occlusion (blood pressure cuff attached to the upper arm inflated to 200 mm Hg or 50 mm Hg higher than the systolic pressure) to determine flow-mediated dilation, a non-invasive measure of conduit artery endothelial-dependent vasodilation. Doppler analysis was used to evaluate mean flow velocity following cuff occlusion, a measure of microvascular function. All vascular images were analyzed at Boston University, using Vascular Research Tools Brachial Analyzer for Research V.6.8.5 (Medical Imaging Applications, IA) by a technician blinded to tobacco product use group. Vascular function was tested at baseline and 30 minutes after acute tobacco product use. Blood pressure and heart rate were measured at baseline and 10 min after use.

### Measurements of urinary metabolites of nicotine and volatile organic compounds

Urine was collected at baseline and 1 h after tobacco product use. Spot-urine collection was used to quantify nicotine and cotinine, a measure of nicotine exposure, at baseline and at 1 h after the acute exposure, by mass spectrometry.^13^ Urinary creatinine levels were measured using Infinity Creatinine Reagent (Thermo Fisher Scientific, MA) on a COBAS MIRA-plus analyzer (Roche, NJ). Samples with levels below the level of detection (LOD) (14.98 ng/mL for nicotine and 2.98 for cotinine) were assigned the value of the LOD/the square root of 2. Metabolites of tobacco-induced aldehydes and other volatile organic compounds were evaluated in the urinary samples using mass spectrometry as previously described.^14, 15^ We measured 22 VOC urinary metabolites (Supplementary Table 1) and evaluated the association with vascular effects of the 11 VOCs with more than 50% of the participants having levels above the LOD: CEMA, 3HPMA, AAMA, CYMA, DHBMA, HPMMA, PGA, 2HPMA, MA, BMA, 3,4 MHA.

### Statistical Analyses

We used one-way ANOVA or chi-squared testing for continuous or categorical data, respectively to compare clinical characteristics and baseline vascular measures across the three groups (pod-based e-cigarette users, combustible cigarette users and non-users). Post-use urinary nicotine, cotinine, and VOCs were natural log transformed given non-normal distribution. Nicotine and cotinine were compared with a general linear model adjusting for urinary creatinine. We used a repeated measures ANOVA to compare the change in vascular measures before and after product use between the three study groups based on an interaction between study group and the time of measurement. Post-hoc analyses to compare changes between individual groups were performed with Tukey adjustment. For vascular measure changes that differed across groups, a general linear model was constructed adjusting for age, sex, race, and site. Association between the post-use urinary VOC levels (after natural log transformation) and the change in flow-mediated dilation, systolic blood pressure, and heart rate were compared by using linear regression models adjusting for urinary creatinine, age, and sex. Data are reported as mean ± standard deviation unless noted. Two-sided p˂0.05 was considered statistically significant. All statistical analyses were carried out using SPSS 24 (IBM, Armonk, NY).

## RESULTS

### Participant product use patterns

Pod-based e-cigarette users were younger than non-users and users of combustible cigarette users (**Table 1**). A similar proportion of women were in the pod-based use group compared with combustible cigarette users, but lower than non-users. Urinary nicotine and cotinine levels were higher in pod-based e-cigarette users and combustible cigarette users than non-users (**Table 1**). Among pod-based users (**Table 2**), 42% were dual users, 27% were exclusive e-cigarette users who were former smokers, and 31% were exclusive users who never used combustible cigarettes. Overall 62% of the pod-based users used JUUL products. Other products included BLU pods, Aspire, Smok, VUSE, Puff Bar, Relx, Suorin, and X1 Smoke. Mint flavors were the most commonly used across the 3 groups. Tobacco flavors were rarely used in the pod-based users who never used combustible cigarettes.

**Table 1.** Clinical Characteristics

|  | Pod-Based<br>Users<br>N=48 | Combustible<br>Cig Users<br>N=21 | Non-Users<br>N=37 | P Value |
| --- | --- | --- | --- | --- |
| **Clinical characteristics** |  |  |  |  |
| Age | 24 ± 5 | 34 ± 7 | 27 ± 5 | <0.0001 |
| Female, N (%) | 16 (33) | 8 (38) | 30 (81) | <0.0001 |
| Race, N |  |  |  | 0.15 |
| American Indian | 0 | 0 | 1 |  |
| Asian | 7 | 2 | 7 |  |
| Black/African American | 5 | 5 | 0 |  |
| White | 31 | 12 | 27 |  |
| Other | 5 | 2 | 2 |  |
| Ln Urinary Cotinine* | 5.2 ± 0.2 | 6.6 ± 0.3 | 0.9 ± 0.3 | <0.0001 |
| Ln Urinary Nicotine* | 4.6 ± 0.2 | 5.1 ± 0.3 | 2.9 ± 0.2 | <0.0001 |
| Ln Urinary Anabasine* | 1.38 ± 0.04 | 1.61 ± 0.06 | 1.33 ± 0.04 | 0.001 |
| Ln Urinary Anatabine* | 0.19 ± 0.09 | 0.93 ± 0.15 | 0.08 ± 0.11 | <0.0001 |
| Mean ± SD or percent as appropriate. |  |  |  |  |
| Cig = cigarette, bpm = beats per min |  |  |  |  |
| *adjusted for urine creatinine, Mean ± SEM |  |  |  |  |

**Table 2.** Characteristics across groups of pod-based e-cigarette users

|  | Dual Users<br>N=20 | Former<br>Smokers<br>N=13 | Never<br>Smokers<br>N=15 | P Value |
| --- | --- | --- | --- | --- |
| **Clinical characteristics** |  |  |  |  |
| Age | 23 ± 4 | 27 ± 6 | 21 ± 2 | 0.001 |
| Female, N (%) | 8 (40) | 2 (15) | 6 (40) | 0.27 |
| JUUL pods, N(%) | 11 (55) | 10 (77) | 9 (60) | 0.28 |
| Flavor Type, N(%) |  |  |  | 0.63 |
| Mint | 9 (45) | 7 (54) | 9 (60) |  |
| Tobacco | 3 (15) | 3 (23) | 0 (0) |  |
| Fruit | 5 (25) | 2 (15) | 4 (27) |  |
| Candy | 3 (15) | 1 (8) | 2 (13) |  |
| Mean ± SD or percent as appropriate. |  |  |  |  |

### Pod-based e-cigarette use and baseline vascular measures

In multivariable models adjusting for age and sex, systolic blood pressure was different between the 3 study groups (**Table 3**), with higher levels in pod-based users and combustible cigarette users (to a comparable degree), as compared with non-users. At baseline, heart rate and diastolic blood pressure were not different across the three groups (**Table 3**). Systolic blood pressure was similar across the pod-based user subgroups (adjusted for age and sex): dual use 121±2mmHg, former smoker 116±3mmHg, never smoker 126±3mmHg, P=0.13) and between JUUL users (121±2mmHg) compared with other pod-based products (121±3mmHg, P=0.9).

**Table 3.** Baseline Vascular Health Measures by Tobacco Use Group: Adjusted Analysis

|  | Pod-Based<br>Users<br>N=48 | Combustible<br>Cig Users<br>N=21 | Non-Users<br>N=37 | P Value |
| --- | --- | --- | --- | --- |
| Baseline brachial diameter, mm | 3.7 ± 0.07 | 3.6 ± 0.11 | 3.7 ± 0.07 | 0.98 |
| Flow-mediated dilation, % | 11.3 ± 0.7 | 11.6 ± 1.2 | 11.7 ± 0.7 | 0.95 |
| Hyperemic flow velocity, cm/sec | 11.5 ± 0.9 | 9.5 ± 1.5 | 8.7 ± 1.0 | 0.17 |
| Systolic blood pressure, mmHg | 122 ± 2 | 121 ± 3 | 112 ± 2 | 0.002 |
| Diastolic blood pressure, mmHg | 74 ± 1 | 75 ± 2 | 70 ± 1 | 0.11 |
| Heart rate, bpm | 66 ± 1 | 65 ± 2 | 63 ± 2 | 0.44 |
| Adjusted for Age and Sex. Least Square Mean ± SEM |  |  |  |  |
| Cig = cigarette, bpm = beats per min |  |  |  |  |

### Acute pod-based e-cigarette use and vascular measures

Following acute use, urinary nicotine and cotinine levels were higher in pod-based e-cigarette and combustible cigarette users than in non-users (**Figure 1**). Following acute use, flow-mediated dilation declined in the pod-based use group compared with the non-use group (**Figure 2**). The decrease in flow-mediated dilation with pod-based e-cigarette use was similar in magnitude to that observed after the use of combustible cigarettes. The difference between the groups remained robust after adjusting for age, sex, race, and study site. The change in flow-mediated dilation was similar across pod-based users who were dual users, former smokers, or never smokers, respectively (−2.9±2.7%, -3.1±3.3%, -3.6±2.5%, P=0.79) and between JUUL users and other pod-based e-cigarette users, respectively (−3.2±3.0%, - 3.1±2.3%, P=0.88). In the pod-based cigarette users, no differences were observed in the change in flow-mediated dilation across the flavor categories or by sex. The post-use nicotine and cotinine levels were associated with the change in flow-mediated dilation (β=-0.69±0.22, P=0.003 and β=0.-44±0.14, P=0.004, respectively), in models adjusting for age, sex, and urinary creatinine. The associations were similar in sensitivity analyses excluding combustible cigarette users.

**Figure 1.**
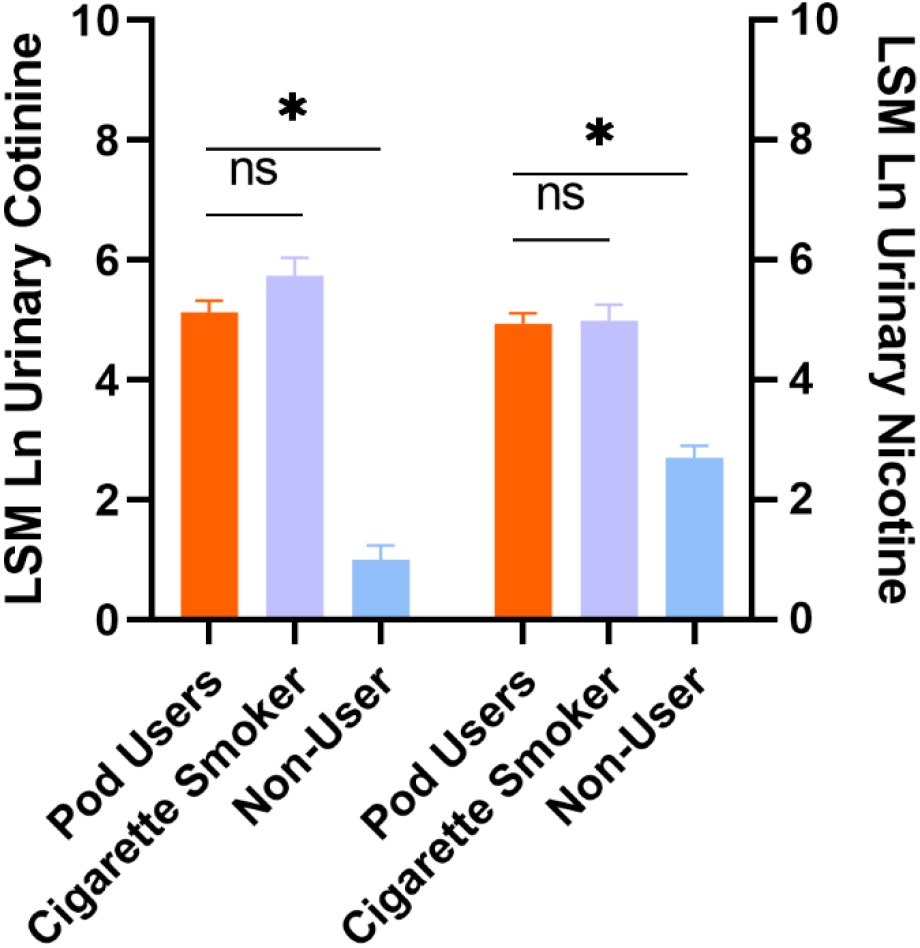
Urinary cotinine (left) and nicotine (right) levels are higher in pod users and combustible cigarette smokers. Compared to non-users, post-use urinary cotinine and nicotine levels were higher in pod users (1.8x10^−25^, 1.7x10^−12^) and combustible cigarette users (1.5x10^−21^, 1.2x10^−8^) by general linear model adjusting for age, sex, and urinary creatinine. Data shown as LSM±SE

**Figure 2.**
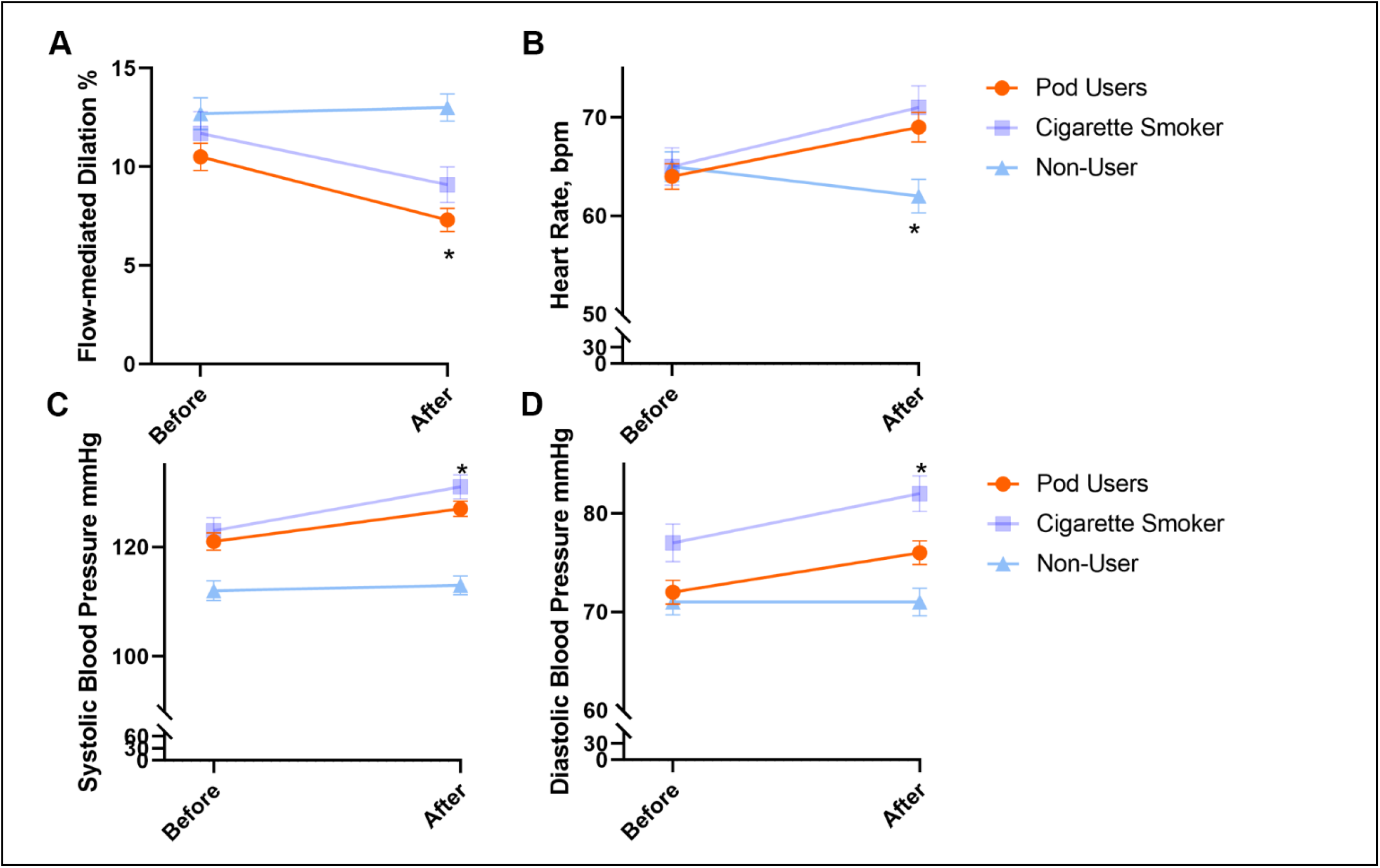
Acute pod-based e-cigarette use lowers vasodilator, raises heart rate and blood pressure. A: The change in flow-mediated dilation following acute use differs across the types of tobacco product used (repeated Measures ANOVA, *P=0.0003 for interaction, Tukey post hoc pod-based vs. non-user P=0.00009, combustible cigarette vs. non-user, P=0.03, pod-based vs. combustible cigarette user, P=0.83). B: The change in heart rate following acute use differs across the types of tobacco product used (*P=1.8x10^−8^, Tukey post hoc pod-based vs. non-user P=2.6x10^−7^, combustible cigarette vs. non-user, P=4.9x10^−7^, pod-based vs. combustible cigarette user, P=0.56). C: The change in systolic blood pressure following acute use differs across the types of tobacco product used (*P=0.00006, Tukey post hoc pod-based vs. non-user P=0.000003, combustible cigarette vs. non-user, P=0.000002, pod-based vs. combustible cigarette user, P=0.45). D: The change in diastolic blood pressure following acute use differs across the types of tobacco product used (*P=0.0001, Tukey post hoc pod-based vs. non-user P=0.003, combustible cigarette vs. non-user, P=0.0003, pod-based vs. combustible cigarette user, P=0.38). Data are expressed as mean ± standard error.

Systolic blood pressure, diastolic blood pressure, and heart rate increased following acute use in the pod-based e-cigarette use group compared with non-use group. The increases following pod-based e-cigarette use were similar to that observed with the use of combustible cigarettes (**Figure 2**). The difference between the groups remained robust in models adjusted for age, sex, race, and study site. The changes in systolic blood pressure, diastolic blood pressure, and heart rate were similar across pod-based users who were dual users, former smokers, or never smokers and between JUUL users and other pod-based e-cigarette users. No differences were observed in the changes in systolic blood pressure and heart rate among pod-based e-cigarette users across different flavors. Post-use nicotine and cotinine levels were associated with the change in systolic blood pressure (β=1.8±0.4, P=0.00008 and β=0.9±0.3, P=0.004), diastolic blood pressure (β=1.2±0.3, P=0.0003 and β=0.9±0.2, P=0.0001), and heart rate (β=2.5±0.4, P=1.1x10^−9^ and β=1.6±0.3, P=8.1x10^−10^), in models adjusting for age, sex, and urinary creatinine. The associations were similar in sensitivity analyses excluding combustible cigarette users.

### Relation of VOC Exposure to Vascular Measures

As shown in **Figure 3**, metabolites of acrylamide (AAMA), acrylonitrile (CYMA), and crotonaldehyde (HPMMA) were consistently associated with greater changes in flow-mediated dilation, systolic blood pressure, and heart rate with acute use. In addition, metabolites of acrolein and xylene were associated with a greater reduction of flow-mediated dilation and increase in systolic blood pressure. Other VOC-derived metabolites were not associated with the changes in vascular measures (**Table 4**). In sensitivity analyses excluding combustible cigarette smokers, the VOC-derived metabolite associations persisted for flow-mediated dilation with 3HPMA, CYMA, and HPMMA; systolic and diastolic blood pressure with AAMA; and heart rate with AAMA and CYMA.

**Figure 3.**
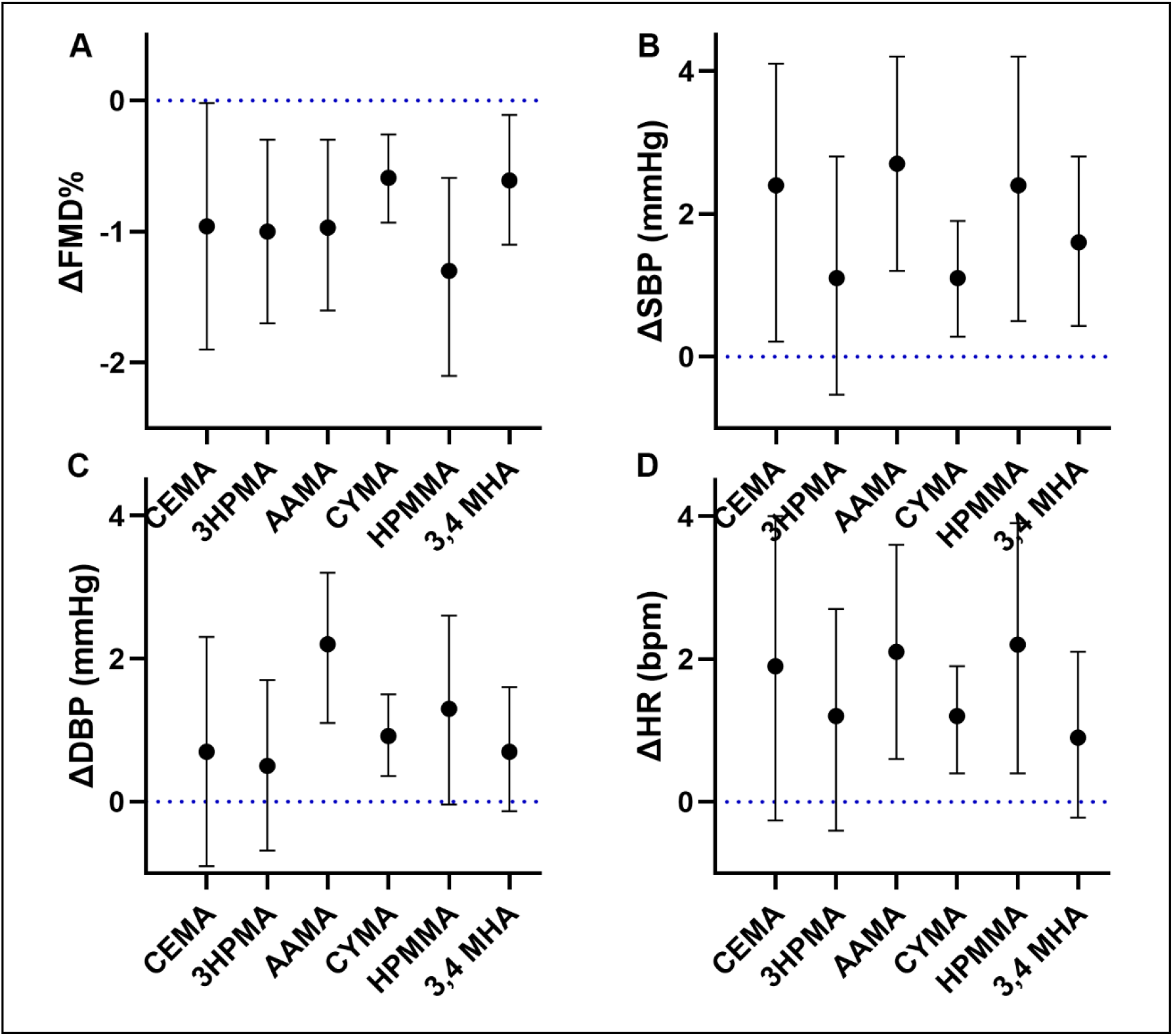
VOC-derived metabolites associated with acute changes in vascular measures. Beta-coefficients and 95% confidence intervals in change in flow-mediated dilation (A), heart rate (B), systolic blood pressure (C), and diastolic blood pressure (D) for each VOC metabolite adjusted for age, sex, and urine creatinine.

**Table 4.** Volatile Organic Compounds and Changes in Vascular Health Measures

| | | ΔFMD | | ΔSBP | | ΔDBP | | ΔHR | |
| --- | --- | --- | --- | --- | --- | --- | --- | --- | --- |
| VOC | Metabolite | Partial<br>r | P | Partial<br>r | P | Partial<br>r | P | Partial<br>r | P |
| Acrolein | CEMA | -0.21 | 0.04 | 0.21 | 0.03 | 0.08 | 0.39 | 0.17 | 0.08 |
| Acrolein | 3HPMA | -0.29 | 0.004 | 0.13 | 0.18 | 0.08 | 0.40 | 0.15 | 0.14 |
| Acrylamide | AAMA | -0.29 | 0.004 | 0.33 | 0.001 | 0.37 | 0.0001 | 0.27 | 0.007 |
| Acrylonitrile | CYMA | -0.34 | 0.001 | 0.26 | 0.009 | 0.31 | 0.002 | 0.30 | 0.002 |
| 1,3-Butadiene | DHBMA | 0.05 | 0.62 | 0.11 | 0.26 | -0.07 | 0.47 | 0.03 | 0.79 |
| Crotonaldehyde | HPMMA | -0.33 | 0.001 | 0.24 | 0.01 | 0.19 | 0.06 | 0.24 | 0.02 |
| Ethylbenzene,<br>styrene | PGA | -0.13 | 0.21 | 0.12 | 0.21 | 0.02 | 0.84 | 0.09 | 0.38 |
| Propylene oxide | 2HPMA | -0.16 | 0.12 | 0.16 | 0.10 | 0.12 | 0.21 | 0.13 | 0.18 |
| Styrene | MA | -0.06 | 0.53 | -0.09 | 0.33 | -0.12 | 0.23 | -0.03 | 0.79 |
| Toluene | BMA | -0.11 | 0.28 | 0.03 | 0.79 | 0.03 | 0.73 | 0.13 | 0.20 |
| Xylene | 3,4MHA | -0.25 | 0.02 | 0.26 | 0.008 | 0.17 | 0.09 | 0.16 | 0.11 |

Adjusted for urinary creatinine, age, sex
VOC-derived metabolites natural log (ln) transformed for analysis

## DISCUSSION

Our study evaluated vascular function in pod-based e-cigarette users in comparison with combustible cigarette users and non-users both at baseline and following acute product use. Our results indicate that at following a tobacco product fast period systolic blood pressure was higher in pod-based e-cigarette users compared with non-users, suggesting sustained alterations of vascular health with regular use. Following acute exposure, pod-based e-cigarette use worsened vasodilator function to a comparable degree compared with that of combustible cigarette use. Similarly, pod-based e-cigarette use induced acute increases in blood pressure and heart rate to an extent comparable with the increases observed in those who smoked a combustible cigarette. The effects of pod-based e-cigarette use on cardiovascular health measures were similar in exclusive users who had never used combustible cigarettes, suggesting that pod-based e-cigarette use affects vascular function even in young adults who use only e-cigarettes. Levels of selected VOC-derived metabolites reflecting exposure to acrolein, acrylamide, acrylonitrile, crotonoaldehyde, and xylene were associated with the changes in vascular function. Overall these findings suggest that the adverse vascular effects of pod-based e-cigarettes are comparable to combustible cigarette smoking, even in young, healthy exclusive e-cigarette users.

Prior studies have reported acute changes in endothelial function in individuals using previous generations of e-cigarettes.^16-18^ One study found that both in chronic users and never users, acute e-cigarette use reduced flow-mediated dilation yet only to a slightly lesser degree than acute combustible cigarette use.^19^ We have previously shown differences in arterial stiffness but not baseline endothelial function in users of earlier generation e-cigarettes who were former smokers.^20^ In another study, no effect was found on microvascular endothelial function following the use of an e-cigarette without nicotine.^21^ A recent study of cigarette smokers who switched to e-cigarettes, with or without nicotine, for one month reported an improvement in resting endothelial function, which might suggest that e-cigarettes could ameliorate some of the effects of smoking.^22^ In a recent acute use study, whereas combustible cigarette use decreased flow-mediated dilation, acute e-cigarette use did not impair vascular function.^23^ The e-cigarette used in this study was a second generation device with only 10 subjects using pod-based e-cigarette.

In a recent animal model study, flow-mediated dilation decreased following exposure to JUUL aerosol to an extent similar to that induced by exposure to cigarette smoke.^8^ A number of prior studies have reported the association of blood pressure and heart rate with non-JUUL e-cigarette use. The preponderance of prior studies have shown an acute increase in blood pressure as well as heart rate following e-cigarette use largely attributable to nicotine exposure.^17, 23^ In a study of cigarette smokers with hypertension switching to e-cigarettes, a reduction in blood pressure has been reported.^24^

Our findings build upon prior work by investigating the effects of pod-based e-cigarettes on measures of vascular function. Unlike prior studies of e-cigarette users who were either current or former combustible cigarette users, our study includes a group of exclusive e-cigarette users who had never used combustible cigarettes. Similar to prior studies, we did not observe a difference in flow-mediated dilation prior to the acute use period suggesting that there may be a reversibility of the vascular effects following a period of tobacco product fasting. In contrast to prior human studies with earlier generations of e-cigarettes, we observed a similar adverse effect of acute pod-based e-cigarette use on endothelium-dependent vasodilation as with combustible cigarettes.^23^ The differences between the two studies may reflect differences in constituents of pod-based e-cigarettes and efficiency of delivery as compared with earlier generation e-cigarettes.^8^ In addition to nicotine, emerging evidence suggests that other components, including flavorings, have endothelial effects.^25, 26^ Differences in study design also may contribute to differential results. In the present study, we evaluated the effect of pod-based e-cigarette use only in e-cigarette users and the effects of combustible cigarette only in exclusive smokers; thus the acute effects may reflect underlying vulnerability due to chronic use. We did not observe differences between JUUL pods or other pod-based devices suggesting that the type of pod device does not alter the effect on vascular function. Further, we found similar effects in young pod-based e-cigarette user who never used combustible cigarettes, which may be important in understanding the implications of the growing numbers of youth adopting exclusive pod-based e-cigarette use.

Our findings of an increase in blood pressure and heart rate with pod-based use suggest that vaping pod-based e-cigarettes results in similar acute hemodynamic effects as earlier generation devices. One important driver of the hemodynamic changes observed with e-cigarette use is likely nicotine.^23, 27^ We observed post-use cotinine and nicotine following pod-based e-cigarette use similar to combustible cigarette use, which is consistent with the possibility that efficient delivery of nicotine with pod-based e-cigarettes accounts for the observed vascular changes. Further work is required to dissociate the effects of nicotine from those due to flavors or or e-liquid solvents propylene glycol and vegetable glycerol, which alone can generate many VOCs in aerosols that induce endothelial dysfunction in animal exposure studies.^28-30^ The association of pod-based e-cigarette use with higher resting blood pressure even following a period of non-use suggests persistent impacts on blood pressure regulation in young adults.

Our findings suggest an effect of selected VOCs on the vascular effects of pod-based e-cigarette use. Prior studies have suggested that e-cigarette use does create to exposure potentially harmful VOCs though it may be lower than with the use of combustible cigarettes.^14, 31, 32^ The association of acrolein metabolites with the changes in vasodilator function is consistent with experimental evidence in animal and cell culture models indicating endothelial injury with acrolein exposure.^28, 33, 34^ In addition, our finding that AAMA levels were associated with all vascular health effects is consistent with a relevance of acrylamide exposure with e-cigarette use.^31^ Taken together, our observations support the value of measuring VOCs when assessing the cardiovascular health implications of novel tobacco products.

The study has a number of limitations that are important to note. The study design required that each participant use their own tobacco product; therefore, it is not possible to isolate the effects of individual constituents such as nicotine, vehicle or flavors. Regarding flavorings, a majority of pod-based users vaped mint-flavored pods, and we did not observe differences across the flavor-types, though larger studies may be needed to detect small differences attributable to different pod flavorings. Each individual in the study used only one product, raising the possibility that differences in characteristics between study participants may contribute to the observed vascular differences. However, the differences did persist in adjusted models and the inclusion of only young, otherwise healthy adults may limit the degree of confounding.

Additionally, there were differences in sex distribution between non-user compared to tobacco use groups, which could contribute to residual confounding. These limitations are balanced by the strengths, including an acute use observation and the consistency of findings across flavors as well as in exclusive pod-based e-cigarette users.

In conclusion, our study provides additional insights into the effects of pod-based e-cigarette use on measures of cardiovascular health in young, healthy adults. The uptake of pod-based e-cigarette vaping in youth and in young adults who are never smokers is of public health concern; thus, it is critical to understand the early cardiovascular implications of pod-based e-cigarettes in youth.^3^ Significantly, we found that pod-based e-cigarette use is associated with acute changes in measures of vascular health, as well as persistent abnormalities in systemic blood pressure as suggested by elevated resting blood pressure at baseline to similar levels in combustible cigarette users. Selected VOC-derived metabolites associate with the vascular health changes suggesting potential toxicity pathways of vascular injury with tobacco product use. Longitudinal studies are needed to evaluate whether the alterations in vascular health translate to earlier development of clinically hypertension, calculated cardiovascular risk or ultimately cardiovascular events. Regulation of pod-based electronic cigarettes needs to take into account the cardiovascular health effects in young adults who were never used combustible cigarette when balancing with for the potential of harm reduction in adult combustible cigarette users. A regulatory framework that assesses the production of toxic VOCs by specific tobacco products and devices may be a feasible approach to reduce cardiovascular injury.

## Data Availability

All data produced in the present study are available upon reasonable request to the authors.

---

**Supplementary Table 1.** Volatile Organic Compounds and their Abbreviations

| **VOC urinary metabolites** | **Abbreviation** |
| --- | --- |
| N-Acetyl-S-(2-carboxyethyl)-L-cysteine | CEMA |
| N-Acetyl-S-(3-hydroxypropyl)-L-cysteine | 3HPMA |
| N-Acetyl-S-(2-cyanoethyl)-L-cysteine | CYMA |
| N-Acetyl-S-(2-hydroxyethyl)-L-cysteine | AAMA |
| t,t-Muconic Acid | MU |
| N-Acetyl-S-(n-propyl)-L-cysteine | BPMA |
| N-Acetyl-S-(3,4-dihydroxybutyl)-L-cysteine | DHBMA |
| N-Acetyl-S-(1-hydroxymethyl-2-propenyl)-L-cysteine | MHBMA1 |
| N-Acetyl-S-(4-hydroxy-2-buten-1-yl)-L-cysteine | MHBMA3 |
| N-Acetyl-S-(3-hydroxypropyl-1-methyl)-L-cysteine | HPMMA |
| N-Acetyl-S-(N-methylcarbamoyl)-L-cysteine | AMCC |
| Phenylglyoxylic acid | PGA |
| N-Acetyl-S-(2-hydroxypropyl)-L-cysteine | 2HPMA |
| N-Acetyl-S-(1-phenyl-2-hydroxyethyl)-L-cysteine +<br>N-Acetyl-S-(2-phenyl-2-hydroxyethyl)-L-cysteine | PHEMA |
| Mandelic Acid | MA |
| N-Acetyl-S-(trichlorovinyl)-L-cysteine | TCVMA |
| N-Acetyl-S-(benzyl)-L-cysteine | BMA |
| N-Acetyl-S-(1,2-dichlorovinyl)-L-cysteine | 12DCVMA |
| N-Acetyl-S-(2,2-dichlorovinyl)-L-cysteine | 22DCVMA |
| Urinary N-Acetyl-S-(dimethylphenyl)-L-cysteine | DPMA |
| 2-Methylhippuric acid | 2MHA |
| 3-Methylhippuric acid + 4-Methylhippuric acid | 3,4MHA |

